# Prevalence of Bacteriologically-Confirmed Tuberculosis in Urban Blantyre, Malawi 2019-20: Substantial Decline Compared to 2013-14 National Survey

**DOI:** 10.1101/2023.04.20.23288872

**Authors:** Helena R A Feasey, McEwen Khundi, Rebecca Nzawa Soko, Emily Nightingale, Rachael M Burke, Marc Y R Henrion, Mphatso D Phiri, Helen E Burchett, Lingstone Chiume, Marriott Nliwasa, Hussein H Twabi, James A Mpunga, Peter MacPherson, Elizabeth L Corbett

## Abstract

Recent evidence shows rapidly changing tuberculosis (TB) epidemiology in Southern and Eastern Africa, with need for subdistrict prevalence estimates to guide targeted interventions. We conducted a TB prevalence survey to estimate current TB burden in Blantyre city, Malawi. From May 2019 to March 2020, 215 households in middle/high-density residential Blantyre, were randomly-selected from each of 72 clusters. Consenting eligible participants (household residents ≥ 18 years) were interviewed, including for cough (any duration), and offered HIV testing and chest X-ray; participants with cough and/or abnormal X-ray provided two sputum samples for microscopy, Xpert MTB/Rif and mycobacterial culture. TB disease prevalence and risk factors for prevalent TB were calculated using complete case analysis, multiple imputation, and inverse probability weighting. Of 20,899 eligible adults, 15,897 (76%) were interviewed, 13,490/15,897 (85%) had X-ray, and 1,120/1,395 (80%) sputum-eligible participants produced at least one specimen, giving 15,318 complete cases (5,895, 38% men). 29/15,318 had bacteriologically-confirmed TB (189 per 100,000 complete case (cc) / 150 per 100,000 with inverse weighting (iw)). Men had higher burden (cc: 305 [95% CI:144-645] per 100,000) than women (cc: 117 [95% CI:65-211] per 100,000): cc adjusted odds ratio (aOR) 2.70 (1.26-5.78). Other significant risk factors for prevalent TB on complete case analysis were working age (25-49 years) and previous TB treatment, but not HIV status. Multivariable analysis of imputed data was limited by small numbers, but previous TB and age group 25-49 years remained significantly associated with higher TB prevalence.

TB prevalence for Blantyre was considerably lower than the 1,014 per 100,000 for urban Malawi in the 2013-14 national survey, at 150-189 per 100,000 adults, but some groups, notably men, remain disproportionately affected. TB case-finding is still needed for TB elimination in Blantyre, and similar urban centres, but should focus on reaching the highest risk groups, such as older men.

## Introduction

The high tuberculosis (TB) incidence in Southern and Eastern African countries has been driven by generalised HIV epidemics [1], together with poverty and urbanisation. In the WHO African Region, TB incidence declined by 22% between 2015 and 2021 [2], concurrent with continuing decreases in HIV incidence and HIV-related deaths [3, 4]. In Malawi, estimated TB incidence has declined from 338 per 100,000 in 2010 to 132 per 100,000 in 2021 [2]. The most recent national TB prevalence survey conducted in 2013-14 estimated an urban prevalence of bacteriologically-confirmed pulmonary TB of 1,014 per 100,000 adults (15+ years) [5, 6] (including Blantyre, the second largest city in Malawi), prompting government and donor investment in TB case-finding activities focused on urban areas. In this time of rapidly declining estimated TB incidence, understanding true TB burden can support more effective National TB Programme (NTP) interventions as countries work towards TB elimination goals TB [7]. As epidemics decline, TB is likely to become more concentrated in marginalised and harder-to-reach groups, requiring increasingly targeted case-finding strategies [8, 9].

TB case-notification rates (CNRs) have been declining in Blantyre [10], likely reflecting the combined effects of increasing coverage with antiretroviral therapy (ART) [11, 12] and use of TB preventive therapy for people living with HIV (PLHIV), as well as TB case-finding and prevention activities. HIV testing and treatment services have been successfully scaled-up to reach UNAIDS 90-90-90 targets for 2020 in Malawi [12, 13]. However, case-notifications are an imperfect guide to TB burden in a population, since they do not include people with TB who remain undiagnosed or may not reach care [2]. As such, low TB CNRs can reflect either under-notification or low incidence of TB. While TB prevalence surveys are laborious and expensive they likely provide the least biased approach to estimating disease burden [14].

The Sustainable Community-based Active case-finding for Lung hEalth (SCALE) trial was designed to investigate the impact of door-to-door active case-finding (ACF) on TB case-notification rates and, if sufficiently powered, the prevalence of undiagnosed TB [15] in Blantyre. In 2019-2020, a TB prevalence survey was, therefore, conducted in all SCALE clusters before the ACF intervention to determine the baseline prevalence of undiagnosed TB and re-evaluate power for intended trial outcomes. The aim was to estimate the burden of TB amongst adults 18 years or older in middle-to-high density urban Blantyre, Malawi.

## Methods

We undertook a cluster-based, cross-sectional TB prevalence survey in middle- to high-density residential areas of Blantyre, Malawi between May 2019 and March 2020. Blantyre City is located in the Southern Region of Malawi, and has a population of approximately 800,250 [16] mostly living in several informal urban settlements built on underserviced land [17]. Although health care is free at the point of care, poorly developed road networks limit access to health and other municipal services. These informal settlements were the focus of this study, which excluded the smaller, central, and more affluent residential and industrial areas. Informal residential areas were demarcated into 72 clusters of approximately 4,400 adult residents each using Community Health Worker catchment area boundaries and population estimates from a city-wide enumeration census conducted with Blantyre District Health Office (DHO) in 2015. The study covered approximately 75% of the geographical area of Blantyre City.

In each cluster, 115 households were randomly selected from a sampling frame of all household GPS co-ordinates obtained from Google Earth, aiming to recruit 215 adults (aged 18 and above) per cluster. Each household was visited at least three times to maximise recruitment of all adult household members, with an initial visit to sensitise household members and book survey appointments. Survey teams covered two clusters per week, with initial activities taking four to five days per cluster, followed by repeat visits to include previously unavailable residents in March 2020.

Household residents were defined as those who usually ate and slept in the same residence. All adult (18 years and over) household residents were eligible for participation if willing and able to provide written or witnessed informed consent. A household questionnaire was conducted with one consenting adult household member (the household head if present) to capture household-level variables including socioeconomic indicators and the age and sex of all household residents. Individual questionnaires were then conducted with all consenting household members, including socio-demographics, a symptom screen for cough of any duration, and brief details of previous HIV and TB testing and care. Participants reporting cough were given two sputum pots, with one collected immediately for microscopy and culture, if possible, and the second collected for Xpert MTB/Rif after an hour or more. All participants were asked to attend a temporary tented digital chest X-ray and HIV testing camp located within each cluster during recruitment days.

Chest X-ray used Min X-ray Commander CMDR-2S-T, with films classed as normal or having any abnormality by a trained radiographer, with reference to results of Qure.ai (version 2) computer-aided detection software. All participants with abnormal X-rays were requested to provide two spot sputum samples. The first sample was taken immediately and the second an hour later. HIV testing used OraQuick (OraSure) and Determine (Alere) finger-prick tests in parallel, with confirmation by Uni-Gold (Trinity Biotech) for positive results, was offered to all participants not on ART. Participants on ART were offered Uni-Gold (Trinity Biotech) confirmatory testing only.

All participants with abnormal X-rays (any abnormality) were referred to an X-ray clinic for review by clinician in a tented community clinic the following week. Those identified as HIV positive and not on ART through the onsite HIV testing were given onsite counselling and referred to the local government HIV clinic for ART initiation.

### Laboratory methods

Sputum samples were processed in the Malawi-Liverpool-Wellcome (MLW)/Kamuzu University of Health Sciences (KUHeS) TB Laboratory, with the first specimen used for fluorescent microscopy (auramine) and liquid culture (Bactec MGIT 960, Becton Dickinson, Franklin Lakes, NJ, USA) and the second specimen used for microscopy and Xpert MTB/Rif (Cepheid, Sunnyvale, CA, USA). Mycobacterial Growth Indicator Tube (MGIT) positive samples were confirmed by Ziehl-Neelsen microscopy (morphology/cording) and MPT64 (SD Bioline, Yongin, Republic of Korea) antigen testing to identify *Mycobacterium tuberculosis (*MTB). Those negative on MPT64 were further incubated at different temperatures on Löwenstein-Jensen (LJ) slopes and classified as non-tuberculous mycobacteria or MTB based on subsequent morphology and growth characteristics.

Participants with positive microscopy, Xpert or MTB culture results were classified as having bacteriologically-confirmed TB. For this survey, an Xpert MTB/Rif G4 trace result was considered positive for MTB. Smear-positive TB participants were defined as those with a direct smear indicating acid fast bacilli. Confirmed TB participants were actively traced for assisted registration for TB treatment at local government clinics.

### Statistical methods

The calculated sample size of 14,511 participants was based on the ability to estimate an overall TB prevalence of 900 per 100,000 with absolute precision of +/- 250,000 per 100,000 (relative precision 27.8%) and a design effect to account for clustering of 2.25. Based on previous work by NTP and our research group in Blantyre, a relatively high non-participation rate of 25% was also assumed. This final sample was rounded up to 15,500 adults (215 per cluster).

Data was summarised by frequencies, percentages, and medians as appropriate, with chi-squared tests to examine differences between groups, such as participation rate by sex.

Following WHO-recommended best-practice analytical methods [18, 19], we estimated TB prevalence using logistic regression models with robust standard errors (calculated from observed between-cluster variability) to account for clustering using three approaches to missing data: 1) complete case analysis (excluding participants eligible for sputum submission but for whom smear, Xpert MTB/Rif and/or culture data were missing); 2) multiple imputation of missing values for sex, age, HIV status, symptom status, X-ray status, sputum results, previous TB, TB contact, crowding and wealth variables, and 3) imputation of missing data for those eligible for sputum submission (cough or abnormal X-ray) with inverse probability weighting to represent all eligible individuals. For multiple imputation of missing values, the predictive mean matching imputation model included all the variables investigated as predictors of bacteriologically-confirmed and smear-positive TB in the multivariable regression model, twenty-five imputed datasets were created, and estimates were combined using Rubin’s rules [20]. Sensitivity analysis was also conducted for an alternate definition of a complete case, restricted to participants with available sputum result data and those who completed both screens.

All analyses were done with R version 4.2.1, using packages including mice [21], lmtest [22] and sandwich [23]. This prevalence survey was part of the SCALE trial with registration number ISRCTN11400592.

#### Data and reproducibility

Data and code to reproduce this analysis is available from *https://osf.io/eu2xf/*.

#### Ethics

The survey protocol was approved by the ethics committee of the London School of Hygiene and Tropical Medicine and Kamuzu University of Health Sciences in Malawi. Written (or witnessed thumbprint if illiterate) informed consent was obtained from all participants.

## Results

Between May 2019 and March 2020, 20,899 eligible adults were enumerated in 7,175 randomly selected and visited households, although many were not physically present during the household visit. 76% (15,897) participated in the survey and underwent symptom screen; 13,490 (85%) had chest X-ray. 1,394/15,897 (9%) participants were eligible to submit sputum through reporting a cough of any duration and/or abnormal X-ray. Of these, 1,140 (82%) submitted at least one sputum sample and 900 submitted two sputum samples (Figure 1).

**Figure 1:**
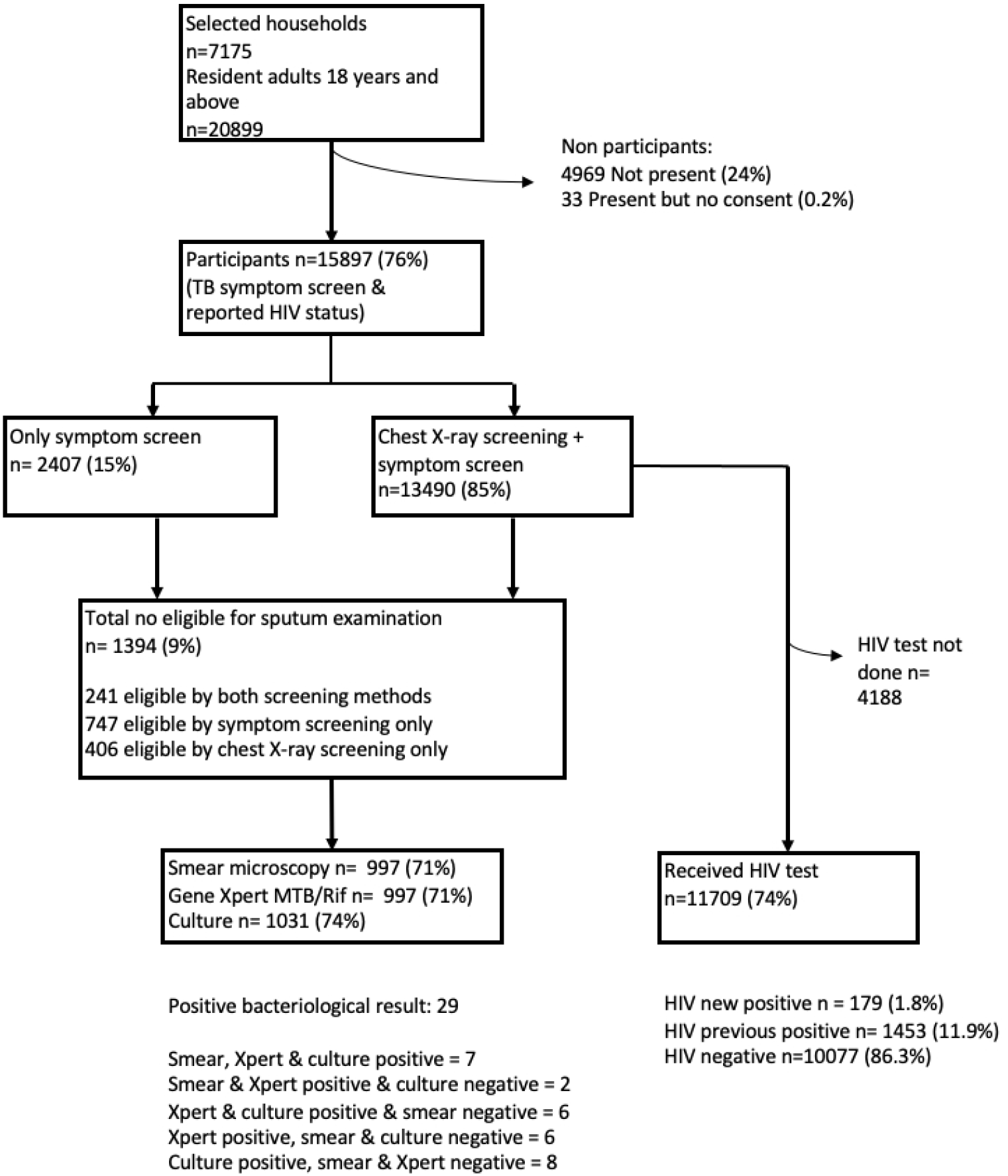
Schematic diagram of number of participants screened for TB & HIV.

Participation rates varied substantially by sex and age group. Participation was higher in women (9,766 of 11,283 [86.6]) than men (6,131 of 9,616 [63.8%], *χ*^2^ p<0.001). The participation rate was highest in people aged 18-24 years (5997 of 7149 [83.9%]) and lower in those aged 25-49 years (8,006 of 10,881 [73.6%]) and 50+ years (1,885 of 2,476 [76.1%], *χ*^2^ p<0.0001). The difference in age participation was driven by men, with the lowest participation rates in working-age men (2,738/4,854 [56.4%] men aged 25-49).

HIV results were available for 11,709 participants of whom 179 (1.8%) were newly identified as HIV-positive, whilst 1,453 (11.9%) were confirmed as previously-known HIV-positive. Overall, 1,971/15,897 (12.4%) participants were identified as HIV-positive, of whom 1,741/1,971 (88.3%) were taking ART.

Previous TB was reported by 456 of 15,897 (2.9%) participants, with 24 (24/15897 [0.2%]) currently on TB treatment; 721 (4.5%) reported knowing someone who had received TB treatment in the last 12-months.

Of the 1,395 participants eligible to submit sputum 1,120 had valid smear results, 1,075 valid culture results, and 900 valid Xpert MTB/Rif results. 579 sputum-eligible participants were missing valid results from at least one sputum tests giving 15,318 complete cases (Table 1). 29 participants were identified with bacteriologically-confirmed TB (one of whom was already on TB treatment). Of those 29, nine were smear-positive (all confirmed by Xpert MTB/Rif or culture) and the others Xpert/culture positive and smear negative (Figure 1 & Supplementary Table 1).

**Table 1:** Prevalence of TB disease per 100,000 adults, with robust standard errors used to calculate 95% confidence intervals.

|  | Total (n) | TB (n) |  | Prevalence (95% CI) |  |
| --- | --- | --- | --- | --- | --- |
|  |  |  | Complete case | Fully imputed | Inverse weighting |
| *All participants* | 15318 | 29 | 189 (132-272) | 139 (71-272) | 150 (76-297) |
| *Sex* |  |  |  |  |  |
| Female | 9423 | 11 | 117 (65-211) | 97 (54-176) | 100 (55-183) |
| Male | 5895 | 18 | 305 (144-645) | 198 (94-415) | 225 (105-479) |
| *Age* |  |  |  |  |  |
| 18-24 | 5811 | 5 | 86 (36-207) | 82 (37-183) | 65 (27-158) |
| 25-49 | 4356 | 19 | 246 (92-657) | 189 (77-468) | 197 (73-531) |
| 50+ | 582 | 5 | 281 (81-966) | 277 (93-821) | 283 (85-938) |
| *HIV status* |  |  |  |  |  |
| HIV negative | 15318 | 23 | 171 (114-257) | 126 (84-189) | 148 (97-228) |
| HIV positive | 5895 | 6 | 320 (130-783) | 270 (116-628) | 292 (122-693) |
| *Previous diagnosis* |  |  |  |  |  |
| No previous TB | 14895 | 25 | 168 (113-248) | 128 (87-189) | 143 (96-214) |
| Previous TB | 423 | 4 | 946 (329-2687) | 613 (214-1742) | 794 (270-2308) |

Notes: HIV status as identified through testing in prevalence survey, or if no test as reported in individual survey 5 of complete cases no age recorded (3 HIV negative, 1 HIV positive & 1 HIV unknown) 626 participants with HIV unknown status but no TB cases amongst them Previous TB includes one currently on TB treatment

Of the 29 participants identified with bacteriologically-confirmed TB, 18 (62%) were male and the highest number were in the 25-49 age group (19 [66%]) (Table 1). In total 6/29 (21%) with bacteriologically-confirmed TB were living with HIV, of whom five were taking ART and one was newly diagnosed. In addition, four (14%) of the 29 identified with prevalent TB reported a previous TB diagnosis (one currently on treatment) and two (7%) reported knowing someone who had started TB treatment in the last 12 months. Fourteen had a cough, of whom nine also had an abnormal chest X-ray. Twelve (41%, 95% CI: 24-61%) had an abnormal chest X-ray but no cough, and for three participants their cough result was missing (two had normal X-ray and one X-ray result was also missing). Overall, of the 29 participants with prevalent bacteriologically-confirmed TB, ten (34%, 95% CI: 18-54%) reported no TB symptoms at household survey (cough, fever, night-sweats or weight-loss) (Supplementary Tables 1 & 2).

The overall prevalence of bacteriologically-confirmed TB was: 189 per 100,000 adults (95% CI 132-272) for the complete case model; 139 per 100,000 adults (95% CI: 71-272) for the multiple imputation model; and 150 per 100,000 adults (95% CI: 76-297) for the inverse probability weighted model (Figure 2). Sensitivity analysis with the alternate complete case definition gave a TB prevalence of 223 per 100,000 adults (95% CI: 155-320).

**Figure 2:**
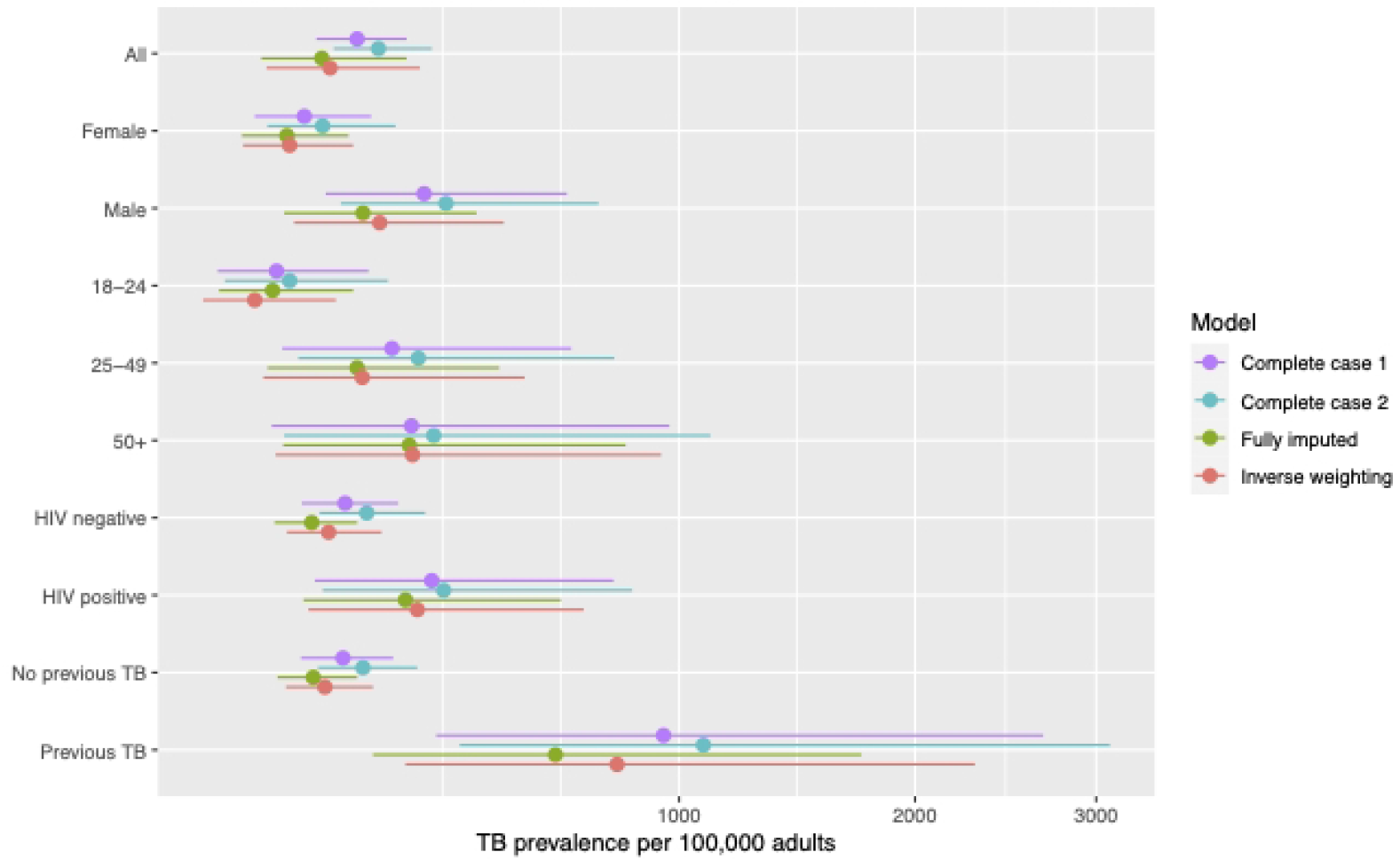
Estimated tuberculosis prevalence by different analytical models. Overall and stratified point prevalence + 95% confidence intervals using robust standard errors

TB prevalence varied considerably by sex, age, and HIV status. The inverse probability weighting model is considered to give the single best estimate of TB prevalence [19] and in our study we considered it most likely to be closest to the true prevalence since it adjusted for the low participation rates amongst working age men (i.e. those aged 25-49 years). In this model male TB prevalence was more than twice as high as female prevalence (225 per 100,000 [95% CI: 105-479] for males vs. 100 per 100,000 [95% 55-183] for females). TB prevalence was highest in the age group 50 years and over (283 per 100,000 [95% CI: 85-938). TB prevalence was also higher in people living with HIV (PLHIV) at 292 per 100,000 (95% CI: 122-693) compared to HIV-negative people (148 per 100,000 [95%CI: 97-228]).

The inverse probability weighting model gave a smear-positive TB prevalence of 37 per 100,000 adults (95% CI: 9-169) (Supplementary Table 3). Smear-positive prevalence was higher for males (84 per 100,000 [95% CI: 18-400]) than females (18 per 100,000 [95%CI: 4-71]), higher amongst PLHIV (132 per 100,000 [95% CI: 33-520]) than those who are HIV negative (42 per 100,000 [95% CI: 20-88]), and highest in the age group 50 years and over (141 per 100,000 [95% CI: 23-852]). The overall highest smear-positive TB prevalence was amongst those who had previously been treated for TB at 349 per 100,000 (95% CI: 74-1640) compared to 42 per 100,000 (95% CI: 21-86) for those with no previous treatment.

On univariable analysis, male sex (OR 2.62, 95% CI: 1.17-6.14, compared to female), being aged 25-49 years (OR 2.86, 95% CI: 1.07-7.68 compared to 18-24 years) and previous TB (OR 5.68, 95% CI: 1.96-16.42, compared to no previous TB treatment were associated with increased odds of prevalent TB (Table 2). On multivariable analysis using the recommended inverse-weighting approach, only previous TB treatment (aOR 3.96, 95% CI: 1.16-13.49) and the age group 25-49 years (aOR 2.69, 95% CI: 1.00-7.26) remained significant predictors of prevalent TB (Table 2). Contact with someone with TB in the last 12 months, male sex, HIV status, crowding and wealth were not significantly associated with prevalent TB in the fully adjusted model. However, this multivariable analysis was limited by the small numbers of participants diagnosed with TB in each category. Due to the small numbers we present age and HIV variables with only three and two categories respectively but analysis with full WHO recommended age and HIV categories is presented in Supplementary Tables 4 and 5.

**Table 2:**
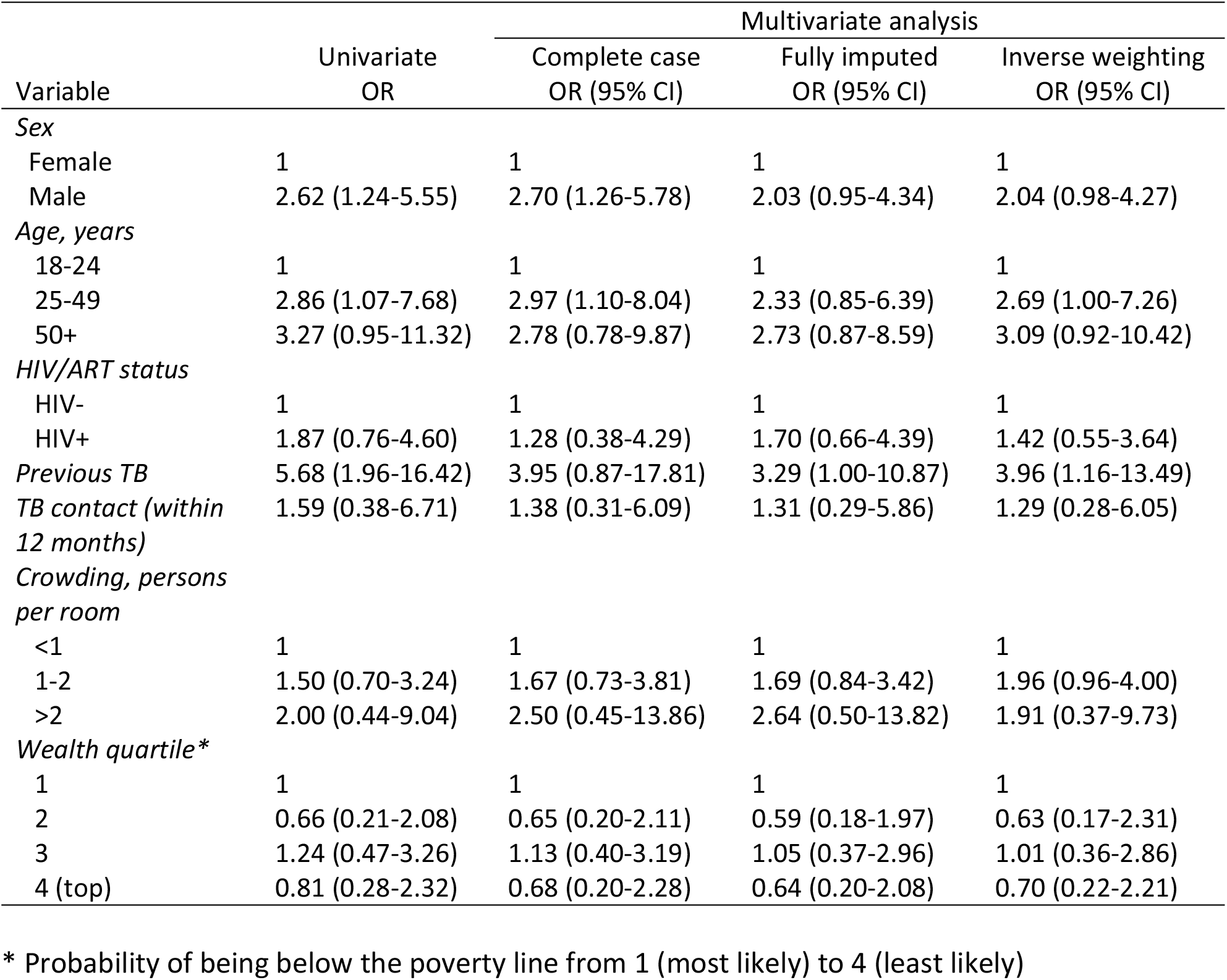
Risk factors for prevalent bacteriologically-confirmed TB, with robust standard errors used to calculate 95% confidence intervals.

| Variable | Univariate<br>OR | Multivariate analysis |  |  |
| --- | --- | --- | --- | --- |
|  |  | Complete case<br>OR (95% CI) | Fully imputed<br>OR (95% CI) | Inverse weighting<br>OR (95% CI) |
| *Sex* |  |  |  |  |
| Female | 1 | 1 | 1 | 1 |
| Male | 2.62 (1.24-5.55) | 2.70 (1.26-5.78) | 2.03 (0.95-4.34) | 2.04 (0.98-4.27) |
| *Age, years* |  |  |  |  |
| 18-24 | 1 | 1 | 1 | 1 |
| 25-49 | 2.86 (1.07-7.68) | 2.97 (1.10-8.04) | 2.33 (0.85-6.39) | 2.69 (1.00-7.26) |
| 50+ | 3.27 (0.95-11.32) | 2.78 (0.78-9.87) | 2.73 (0.87-8.59) | 3.09 (0.92-10.42) |
| *HIV/ART status* |  |  |  |  |
| HIV- | 1 | 1 | 1 | 1 |
| HIV+ | 1.87 (0.76-4.60) | 1.28 (0.38-4.29) | 1.70 (0.66-4.39) | 1.42 (0.55-3.64) |
| *Previous TB* | 5.68 (1.96-16.42) | 3.95 (0.87-17.81) | 3.29 (1.00-10.87) | 3.96 (1.16-13.49) |
| *TB contact (within 12 months)* | 1.59 (0.38-6.71) | 1.38 (0.31-6.09) | 1.31 (0.29-5.86) | 1.29 (0.28-6.05) |
| *Crowding, persons per room* |  |  |  |  |
| <1 | 1 | 1 | 1 | 1 |
| 1-2 | 1.50 (0.70-3.24) | 1.67 (0.73-3.81) | 1.69 (0.84-3.42) | 1.96 (0.96-4.00) |
| >2 | 2.00 (0.44-9.04) | 2.50 (0.45-13.86) | 2.64 (0.50-13.82) | 1.91 (0.37-9.73) |
| *Wealth quartile\** |  |  |  |  |
| 1 | 1 | 1 | 1 | 1 |
| 2 | 0.66 (0.21-2.08) | 0.65 (0.20-2.11) | 0.59 (0.18-1.97) | 0.63 (0.17-2.31) |
| 3 | 1.24 (0.47-3.26) | 1.13 (0.40-3.19) | 1.05 (0.37-2.96) | 1.01 (0.36-2.86) |
| 4 (top) | 0.81 (0.28-2.32) | 0.68 (0.20-2.28) | 0.64 (0.20-2.08) | 0.70 (0.22-2.21) |

\* Probability of being below the poverty line from 1 (most likely) to 4 (least likely)

Significant predictors of prevalent smear-positive TB were male sex (OR 5.35 [95% CI: 1.02-52.74]) and reported previous TB (OR 9.86, 95% CI: 1.00-52.01) on univariable analysis, but no predictors were significant on multivariable analysis, likely due to the small numbers involved (only nine smear-positive TB cases identified).

## Discussion

The main finding from this survey is that the estimated adult prevalence of bacteriologically-confirmed pulmonary TB in Blantyre from this survey – 150 per 100,000 in the inverse probability weighted model – was more than 80% lower than the previous estimates for urban areas in the 2013-14 Malawi National TB Prevalence Survey (1,014 per 100,000 adults aged 15+) [5, 6]. This large decrease in TB prevalence over six years is likely due to both extensive local case-finding efforts and the concurrent rapid scale-up and high coverage of ART for treatment of HIV [12], and potentially TB preventive therapy for PLHIV. As expected [24], TB prevalence was higher amongst men, PLHIV, and those reporting previous TB, as well as among people aged 50 years and over. More than half of survey participants (52%) identified with TB through the survey did not self-report cough, and so would not have been identified through the symptom screen alone, highlighting the benefit of including X-ray or other screening strategies able to identify subclinical TB [25]. As TB epidemics decline in Southern and Eastern Africa, TB disease is likely to become increasingly concentrated in marginalised and hard-to-reach groups requiring adaptive targeted strategies informed by evidence such as local prevalence surveys.

Between the 2013-2014 NTP prevalence survey and this 2019-2020 survey, annual TB case notification rates, including bacteriologically confirmed TB, had been declining in Blantyre [10] with concurrent steep reductions in the percentage of primary care clinic attendees with bacteriologically-confirmed TB following self-presentation for investigation of TB symptoms. [26, 27]. As such, the pronounced decline in the prevalence of undiagnosed TB that we infer from comparing our results to the 2013-14 national survey is almost certainly correct, although our survey still demonstrates under-diagnosis of TB in Blantyre with a prevalence to case-notification ratio of 4.49 (95% CI: 0.98–11.91) as reported elsewhere [10]. Undiagnosed infectious TB remains well above TB elimination targets, underscoring the need to continue appropriately targeted case-finding activities in Blantyre.

Our survey was intended to inform endpoints for a TB case-finding intervention trial, and so used random household sampling in purposively selected study-clusters, and 18+, not 15+, age to define adults as in the 2013-14 National survey. If anything, however, this is likely to over-estimate the municipal burden of undiagnosed TB in Blantyre City compared to National 2013-14 estimates. Our survey area covered most of urban Blantyre, from both geographic and population perspectives. Nationally, Blantyre City has the highest TB case-notifications, reflecting the more densely populated higher HIV-prevalence southern region of Malawi, making our finding even more striking as the 2013-14 urban estimates included cities with lower case-notifications [28, 29].

As expected from the literature [24] and recent national TB prevalence surveys [5, 6], TB prevalence was considerably higher amongst men than women, with the sex ratio increased from 1.33 (95% CI 0.94-1.87) in 2014 to 2.04 (95% CI 0.98-4.27) in this survey, highlighting the need for future case-finding efforts targeted at men. We had limited power to address HIV as a risk factor for undiagnosed TB, with only 6 of 29 (21%) people with prevalent TB being HIV positive, but this does suggest a decrease compared with an estimated 45% of patients diagnosed with TB being HIV-positive in the 2013-14 national TB prevalence survey [30]. This decrease is consistent with more complete diagnosis of HIV, better TB prevention (ART, isoniazid) and better implementation of sensitive (Xpert-based) screening guidelines for patients attending ART clinics in Malawi [7, 31]. It also reflects the estimated 20% decline in HIV prevalence in Blantyre from 17.7% in 2015-16 to 14.2% in 2020-21 [32], and the concurrent improved management of HIV through ART, as evidenced by viral load suppression increasing from 59.5% in 2015-16 [3] to 81.0% in 2021 [32]. Although PLHIV have much higher incidence of TB disease, driving higher case-notifications at facilities, HIV has less impact on undiagnosed prevalent TB due to more rapid progression [33, 34]. In this survey in Blantyre, as in prevalence surveys across much of Africa [35, 36] most patients with undiagnosed infectious TB in the community were HIV-negative (23 out of 29 patients) As TB and HIV prevalence continues to fall, future community case-finding activities will need to be targeted at those at highest risk; for example in this prevalence survey, working age and older men who accounted for over half of all infectious TB patients, and tend to report suboptimal health-seeking [37] placing them at risk of remaining undiagnosed for prolonged periods without detection.

Previous TB treatment and older age groups were associated with higher undiagnosed TB prevalence, but other measured potential risk factors [38] (such as crowding and wealth) were not strongly associated with undiagnosed TB in this study. In part this may reflect our small number of cases. Nevertheless, the proportions of participants with infectious TB who had been previously treated (10% versus 14%) or were currently on TB treatment (3% vs 3%) were strikingly similar in 2019-20 and the 2013-14 National survey, and are consistent with a well-functioning routine treatment programme in Malawi. The age-group at greatest risk of TB (older adults) is also consistent with the 2013-14 National Survey findings, aligns with the known natural history of TB in endemic settings, and may also indicate the “aging” HIV epidemic in this global region [3, 32].

Half (15/29, 52%) of participants identified with TB in our Blantyre survey did not have a cough and would have been missed if only a cough symptom screen was used, and a third (10/29) had none of the WHO four TB symptoms. This aligns with other TB prevalence surveys [25] where typically half of those identified with TB were asymptomatic on the symptom screen. Again this has implications for future case-finding approaches, which should consider intensified screening with highly sensitive tools, such as digital chest X-ray, to identify people with subclinical TB in communities and clinics [39]. However, this may be less applicable in areas with higher TB prevalence since the pronounced declines in undiagnosed TB reported above were achieved in Blantyre with minimal systematic screening for sub-clinical TB.

This study has some limitations, including low precision from the small number of cases in our survey, due to lower than anticipated prevalence, and low rates of participation, particularly amongst working age men. Lower male participation has been seen in nearly all TB prevalence surveys [35], and future surveys should explore methods to increase participation such as further community engagement and study sites/times that are accessible to everyone, including working men, to reduce potential bias from under-participation and/or missing data. However, estimates from the use of different analytical models and imputation methods did not vary considerably suggesting bias due to missing data was limited. Two TB patients were diagnosed with TB but with records showing no cough and a normal X-ray, suggesting inaccurate data. Strengths of the survey include offering HIV testing, high rates of sputum submission from those eligible (82% overall and 60% amongst those with no cough but abnormal chest X-ray) [40], linkage to care for those diagnosed with TB and the use of all three sputum tests (smear microscopy, Xpert MTB/Rif and MGIT culture) to ensure high-sensitivity once sputum was submitted.

Our study demonstrates a substantial decrease in TB prevalence in urban Malawi over the eight years before the COVID-19 pandemic. To build on this and reverse any increase due to COVID-19 [41], future case-finding in Blantyre and similar urban centres in sub-Saharan Africa, should target the highest risk groups such as working-age and older men.

## Data Availability

All data is available within the results, supplementary material and online data repository https://osf.io/eu2xf/

## Acknowledgments

We acknowledge Vincent Phiri and George Sinjani for their help in setting up the data capture systems and overseeing fieldwork.

## References

1. Surie D, Borgdorff MW, Cain KP, Click ES, DeCock KM, Yuen CM. Assessing the impact of antiretroviral therapy on tuberculosis notification rates among people with HIV: a descriptive analysis of 23 countries in sub-Saharan Africa, 2010-2015. BMC Infect Dis. 2018;18(1):481.

2. World Health Organization. Global Tuberculosis Report 2022. 2022.

3. PHIA Project. Malawi Population-based HIV Impact Assessment 2015-2016. 2018. Columbia University; 2016.

4. Joshi K, Lessler J, Olawore O, Loevinsohn G, Bushey S, Tobian AAR, et al. Declining HIV incidence in sub-Saharan Africa: a systematic review and meta-analysis of empiric data. J Int AIDS Soc. 2021;24(10):e25818.

5. Ministry of Health National TB Control Programme. Technical report: Malawi tuberculosis prevalence survey (2013–2014). 2016.

6. World Health Organization. National Tuberculosis Prevalence Surveys 2007-2016. 2021.

7. Republic of Malawi Ministry of Health. National Tuberculosis Control Programme - Programme Manual, Eighth Edition. Malawi: Malawi Ministry of Health, Unit CHS; 2018.

8. European Centre for Disease Prevention and Control. Guidance on tuberculosis control in vulnerable and hard-to-reach populations. 2016.

9. Trauer JM, Dodd PJ, Gomes MGM, Gomez GB, Houben R, McBryde ES, et al. The Importance of Heterogeneity to the Epidemiology of Tuberculosis. Clin Infect Dis. 2019;69(1):159–66.

10. Khundi M, Carpenter JR, Corbett EL, Feasey HRA, Soko RN, Nliwasa M, et al. Neighbourhood prevalence-to-notification ratios for adult bacteriologically-confirmed tuberculosis reveals hotspots of underdiagnosis in Blantyre, Malawi. PLoS One. 2022;17(5):e0268749.

11. Burke RM, Henrion MYR, Mallewa J, Masamba L, Kalua T, Khundi M, et al. Incidence of HIV-positive admission and inpatient mortality in Malawi (2012-2019). Aids. 2021;35(13):2191–9.

12. Kanyerere H, Girma B, Mpunga J, Tayler-Smith K, Harries AD, Jahn A, et al. Scale-up of ART in Malawi has reduced case notification rates in HIV-positive and HIV-negative tuberculosis. Public Health Action. 2016;6(4):247–51.

13. Chirwa Z, Kayambo F, Oseni L, Plotkin M, Hiner C, Chitsulo C, et al. Extending beyond Policy: Reaching UNAIDS’ Three “90”s in Malawi. Front Public Health. 2018;6:69.

14. Borgdorff MW. New measurable indicator for tuberculosis case detection. Emerg Infect Dis. 2004;10(9):1523–8.

15. Feasey HRAK, M.; Nzawa Soko, R.; Bottomley, C.; Chiume, L.; Nliwasa, M.; Twabi, H.; Mpunga, J.A.; Fielding, K.; MacPherson, P.; Corbett, E.L.. OA17-324-09 Impact of active TB case-finding on case notifications in Blantyre, Malawi: a community-based cluster-randomised trial (SCALE). The Union World Conference on Lung Health; Virtual 2022.

16. National Statistical Office (NSO). 2018 Malawi Population and Housing Census. Blantyre City Report. 2021.

17. United Nations Human Settlements Programme (UN-HABITAT). Malawi: Blantyre Urban Profile. 2011.

18. World Health Organization. Tuberculosis Prevalence Surveys: a handbook. 2011.

19. Floyd S, Sismanidis C, Yamada N, Daniel R, Lagahid J, Mecatti F, et al. Analysis of tuberculosis prevalence surveys: new guidance on best-practice methods. Emerg Themes Epidemiol. 2013;10(1):10.

20. Rubin DB. Multiple Imputation after 18+ Years. Journal of the American Statistical Association. 1996;91(434):473–89.

21. van Buuren S. G-OK, Vink G. et al.,. mice: Multivariate Imputation by Chained Equations R package 2022 [Available from: https://cran.r-project.org/web/packages/mice/index.html.

22. Hothorn T. ZA, Farebrother R.W. et al.,. lmtest: Testing Linear Regression Models R Package 2022 [Available from: https://cran.r-project.org/web/packages/lmtest/index.html.

23. Zeileis A. LT, Graham N., Koell S.. sandwich: Robust Covariance Matrix Estimators R Package 2022 [Available from: https://cran.rproject.org/web/packages/sandwich/index.html.

24. Horton KC, MacPherson P, Houben RMGJ, White RG, Corbett EL. Sex Differences in Tuberculosis Burden and Notifications in Low- and Middle-Income Countries: A Systematic Review and Meta-analysis. PLoS Medicine. 2016;13(9).

25. Frascella B, Richards AS, Sossen B, Emery JC, Odone A, Law I, et al. Subclinical Tuberculosis Disease—A Review and Analysis of Prevalence Surveys to Inform Definitions, Burden, Associations, and Screening Methodology. Clinical Infectious Diseases. 2021;73(3):e830–e41.

26. MacPherson P, Webb EL, Kamchedzera W, Joekes E, Mjoli G, Lalloo DG, et al. Computer-aided X-ray screening for tuberculosis and HIV testing among adults with cough in Malawi (the PROSPECT study): A randomised trial and cost-effectiveness analysis. PLoS Med. 2021;18(9):e1003752.

27. Divala TH, Fielding KL, Kandulu C, Nliwasa M, Sloan DJ, Gupta-Wright A, et al. Utility of broad-spectrum antibiotics for diagnosing pulmonary tuberculosis in adults: a systematic review and meta-analysis. Lancet Infect Dis. 2020;20(9):1089–98.

28. Nyirenda T. Epidemiology of Tuberculosis in Malawi. Malawi Med J. 2006;18(3):147–59.

29. The World Bank. Policy Brief: Optimizing Investments in the Tuberculosis Response in Blantyre, Lilongwe, and Mzimba Districts, Malawi: Results of a TB Allocative Efficiency Study.. 2020.

30. Data reported by countries to WHO and estimates of tuberculosis burden generated by WHO for the Global Tuberculosis Report [Internet]. 2022 [cited 14/12/2022]. Available from: https://www.who.int/teams/global-tuberculosis-programme/data.

31. World Health Organisation. Guidelines for intensified tuberculosis case-finding and isoniazid preventive therapy for people living with HIV in resource-constrained settings. 2011.

32. PHIA Project. Malawi Population-based HIV Impact Assessment (MPHIA) Summary Sheet 2020-2021. 2022.

33. Kwan CK, Ernst JD. HIV and tuberculosis: a deadly human syndemic. Clin Microbiol Rev. 2011;24(2):351–76.

34. Corbett EL, Charalambous S, Moloi VM, Fielding K, Grant AD, Dye C, et al. Human immunodeficiency virus and the prevalence of undiagnosed tuberculosis in African gold miners. Am J Respir Crit Care Med. 2004;170(6):673–9.

35. Law I, Floyd K. National tuberculosis prevalence surveys in Africa, 2008-2016: an overview of results and lessons learned. Trop Med Int Health. 2020;25(11):1308–27.

36. Marx FM, Hesseling AC, Martinson N, Theron G, Cohen T. National survey in South Africa reveals high tuberculosis prevalence among previously treated people. Lancet Infect Dis. 2022;22(9):1273.

37. Chikovore J, Hart G, Kumwenda M, Chipungu GA, Desmond N, Corbett L. Control, struggle, and emergent masculinities: a qualitative study of men’s care-seeking determinants for chronic cough and tuberculosis symptoms in Blantyre, Malawi. BMC Public Health. 2014;14:1053.

38. Narasimhan P, Wood J, Macintyre CR, Mathai D. Risk factors for tuberculosis. Pulm Med. 2013;2013:828939.

39. World Health Organization. WHO consolidated guidelines on tuberculosis. Module 2: screening – systematic screening for tuberculosis disease. 2021.

40. Habib SS, Asad Zaidi SM, Jamal WZ, Azeemi KS, Khan S, Khowaja S, et al. Genderbased differences in community-wide screening for pulmonary tuberculosis in Karachi, Pakistan: an observational study of 311 732 individuals undergoing screening. Thorax. 2022;77(3):298–9.

41. Soko RN, Burke RM, Feasey HRA, Sibande W, Nliwasa M, Henrion MYR, et al. Effects of Coronavirus Disease Pandemic on Tuberculosis Notifications, Malawi. Emerg Infect Dis. 2021;27(7):1831–9.

